# Waning Immunity and Partial Vaccination Coverage Lead to Transitions in the Source of Daily Incidence

**DOI:** 10.64898/2026.03.12.26348258

**Authors:** Nora Heitzman-Breen, Sabina L. Atlus, jimi adams, Andrea G. Buchwald, Vanja Dukic, Bailey K. Fosdick, Debashis Ghosh, Jon Samet, Elizabeth J. Carlton, David M. Bortz

## Abstract

Vaccine-acquired immunity plays an important role in controlling the spread of many infectious diseases; however, vaccine efficacy can diminish over time. This work uses a mathematical model to study the effects of *waning vaccination-acquired immunity* on infection incidence. With an SEIR-type compartmental model that considers both vaccinated and unvaccinated populations (and their mixing), we present mathematical conditions under which vaccinated individuals drive ongoing growth in infections, i.e., over half of the daily incidence arises from vaccinated individuals. Analysis of a mathematical model of COVID-19 spread in the state of Colorado suggests how and for what duration vaccinated individuals could have sustained such growth. Importantly, our model demonstrates that, despite potential for brief vaccinated-driven periods of growth in infections, which occur among unvaccinated-driven periods of growth in infections, increased vaccination coverage always reduces total cases and total hospitalizations. This work provides insight into how waning immunity in vaccinated populations can contribute to ongoing infection incidence and demonstrates the value of complementary interventions to prevent disease spread in vaccinated populations.

## 1 Introduction

Vaccine-acquired immunity plays an important role in controlling the spread of many infectious diseases. However, while some vaccines are highly effective and require only a single round of vaccination to protect against infection (e.g., small pox, yellow-fever, polio), other vaccines are only temporarily effective, allowing for periodic outbreaks and requiring frequent repeated vaccination. One such example, seasonal influenza, is responsible for 290,000 to 650,000 deaths globally each year [1], and as of August 2024, there have been over 7.1 million deaths globally and 1.1 million deaths within the United States caused by COVID-19 [2]. Furthermore, while influenza and COVID-19 vaccinations reduce both hospitalizations and mortality [3–5], they do not completely inhibit disease spread [3, 4, 6, 7].

Broadly speaking, compartmental epidemiological models are important tools that have been used to model disease spread and public health interventions for a wide range of human-transmitted diseases such as smallpox [8, 9], measles [10], influenza [11, 12], and COVID-19 [13–18]. Standard mathematical analysis of these models reveals that prevention of disease spread through vaccination is strongly impacted by *herd immunity* [19, 20]. Herd immunity is defined as a threshold level of immunity at which disease does not spread in a population because there are insufficient susceptible individuals. The proportion of a population that must maintain vaccine-acquired immunity can be estimated through a conventional Kermack-McKendrick (compartmental) epidemiological modeling as (1 −1/ℛ_0_)/*e* where ℛ_0_ is the population reproduction number and *e* is the effectiveness at preventing infection [20]. If *e* is too low, achieving herd immunity will be impossible. Furthermore, achieving herd immunity in diseases such as influenza or COVID-19 can be inhibited by several processes including waning immunity [4, 7, 21–23], limited immunity in vulnerable populations [5, 24], vaccine evasion by emerging variants [4, 25–28], and variability of vaccine acceptance by the population [29, 30]. Some modeling work has also predicted the possibility of vaccination conditions where reductions in disease severity with imperfect blocking of transmission could facilitate conditions where pathogens causing severe disease can emerge [31]. Therefore, it is valuable to consider designing public health interventions to account for imperfect vaccination in general, and in particular for the effect of waning immunity on infectious disease transmission.

In this study, we aim to characterize the relationship between waning immunity and the daily incidence in a population of both vaccinated and unvaccinated individuals. The central conclusion is that if vaccine-induced immunity wanes sufficiently fast and vaccination coverage is high, the mathematics leads us to conjecture that vaccinated individuals can be a significant driver sustaining new infections after sufficient time has passed, such that vaccine-acquired immunity has waned. To demonstrate this, Figure 1A depicts the solution to a compartmental epidemiology model (from Section 4.1) of a population of 100, 000 people where vaccine-acquired immunity lasts (on average) 213 days, vaccine coverage is 85%, and the vaccine is assumed 100% effective at blocking transmission at administration. While unvaccinated sources drive the initial new infections, the 25-day-long orange region represents the time window during which more than half of the daily incidence arises from vaccinated sources. As detailed below, the parameters in this model are informed by existing epidemic data, and the figure serves to demonstrate that, with imperfect vaccine effectiveness, vaccinated individuals can be a major source of daily incidence as the number of infections grows. Moreover, this has implications for non-pharmaceutical intervention strategies, which we will also discuss.

**Figure 1:**
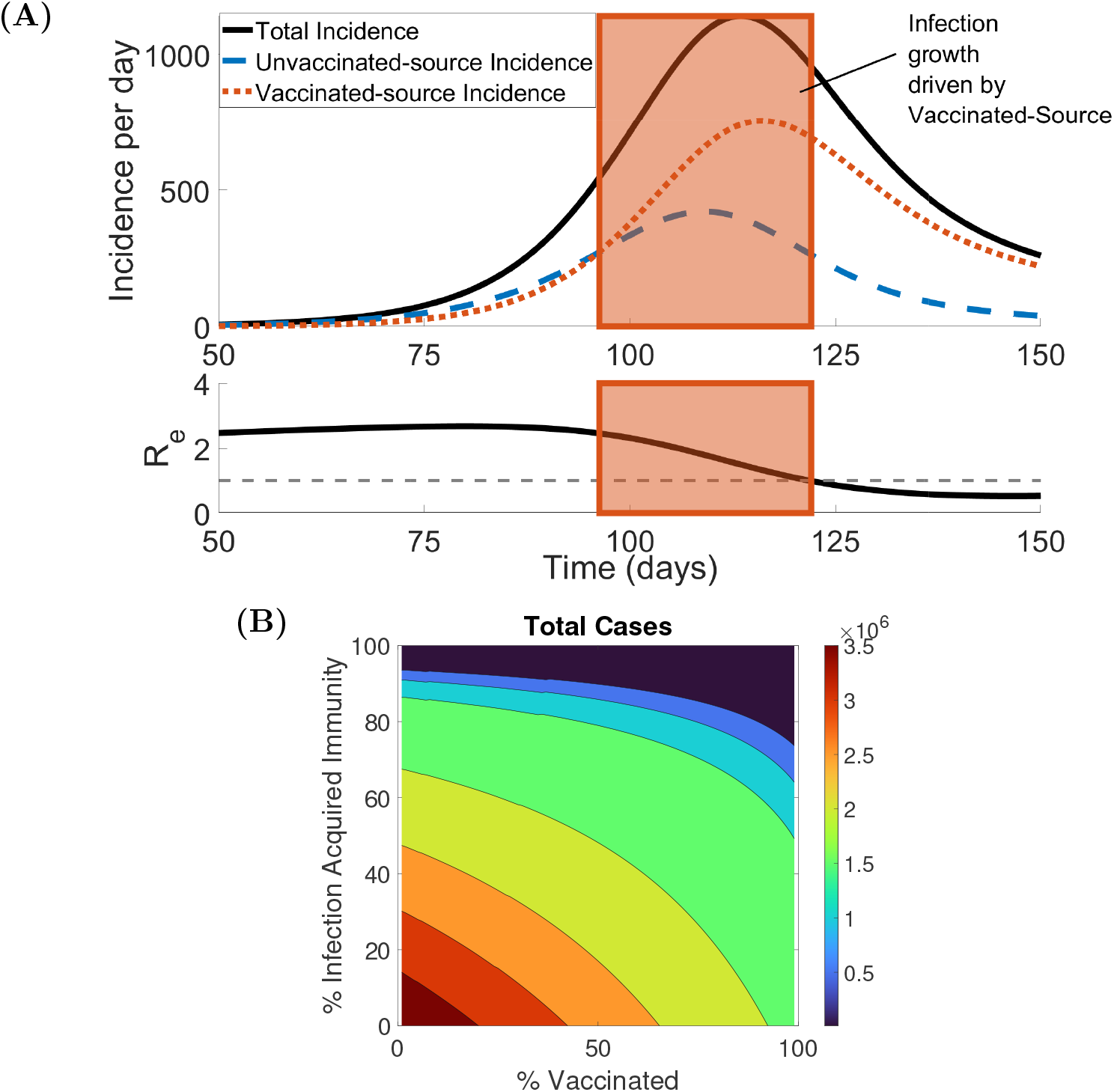
**A. Example trajectory of a compartmental model with waning immunity, where over a 25-day period, over half of the incidence per day is from a vaccinated source.** The solid black line is the total incidence per 100,000 generated from a general compartmental model, Model **G**, using the parameters from Table S1 in the Supplementary Materials. On day 0, we assume that 85% of the population is vaccinated. Incidence is classified as either unvaccinated-source incidence (ℐ_**U**→_), where the infectious individual is unvaccinated (blue dashed line), or vaccinated-source incidence (ℐ_**V**→_), where the infectious individual is vaccinated (red dotted line). Note that the sum of ℐ_**U**→_ and ℐ_**V**→_ is equal to the total incidence per day. The effective reproductive number, ℛ_*e*_, is also plotted over 100 days, and the orange box denotes the 25-day period where vaccinated-source incidence is the majority of incidence per day and ℛ_*e*_ *>* 1. **B. Total cases of an infection are always decreased when either population vaccine-acquired or infection-acquired immunity increases**. Contour plot of the cumulative total cases over 200 days versus the proportion of the population vaccinated and the proportion of the population with infection-acquired immunity generated from Model **G** with parameters from Table S1 in the Supplementary Materials.

In Section 2, we develop a mathematical classification of infection spread based on epidemiological parameters, including an exploration of the impact of levels of population mixing between vaccinated and unvaccinated individuals. In Section 3 we discuss the implications of this work in terms of pharmaceutical and non-pharmaceutical interventions. Lastly, in Section 4, we present compartmental epidemiology models of infection that encapsulate waning immunity. Section 4.1 contains a simplified model to investigate the general idea, while Section 4.2 contains a model of the disease dynamics during the COVID-19 pandemic in the state of Colorado in 2020-2022.

## 2 Results

In this section, we simulate a generalized compartmental transmission model and a Colorado COVID-19-specific transmission model, which account for partial vaccination, vaccine-acquired immunity, waning immunity, and mixing of unvaccinated and vaccinated populations. We observe with the generalized compartmental transmission model, Model **G**, that under specific conditions the incidence from infected vaccinated sources (ℐ_**V**→_) can sustain ongoing growth in infections. We consider three types of infection spread (summarized in Table 1): I) where ℐ_**V**→_ is never the majority of incidence, II) where ℐ_**V**→_ becomes the majority of incidence but not during a period of growth in infections, and III) where ℐ_**V**→_ is the majority of incidence during a period of growth in infections. We determine a partition of the parameter space, for which specific parameter combinations will predict which of the three infection spread types will occur. This partitioning is in terms of unvaccinated and vaccinated population mixing (assortivity), vaccination coverage, and length of vaccine-acquired immunity. Further, we find that the infection spread type impacts the effectiveness of population and sub-population level masking implementation. Lastly, analysis of a Colorado COVID-19 transmission model, Model **C**, suggests that there was a period in 2022 during which vaccinated individuals comprised the majority of incidence during a time of growth in infections.

**Table 1:** Description of infection spread types.

| Infection Spread Type | Description |
| --- | --- |
| Type I | $\mathcal{I}_{\mathbf{V} \rightarrow}$ is never the majority of incidence |
| Type II | $\mathcal{I}_{\mathbf{V} \rightarrow}$ becomes the majority of incidence but not during a period of growth in infections |
| Type III | $\mathcal{I}_{\mathbf{V} \rightarrow}$ is the majority of incidence during a period of growth in infections |

### 2.1 General Transmission Model

The incidence trajectory from Model **G** depicted in Figure 1A demonstrates that it is possible for vaccinated-source incidence (ℐ_**V**→_) to be the majority of incidence during an ongoing period of new infections. However, it is important to note that increasing a population’s vaccination coverage always results in fewer total cases. We studied the effect of different types of initial immunity of the population on total cases during a period of Type III infection spread, where ℐ_**V**→_ is the majority of incidence during a period of growth in infections. We varied the proportion of the population with infection-acquired immunity, *ξ*, and the proportion of the population vaccinated, *ϕ*, while considering the cumulative cases over the first 200 days after the initial case (see Figure 1B). As expected, when population immunity increases, the total case count decreases. While perfect vaccination coverage is desirable, in this work we are interested in transmission dynamics when less than 100% of the population is vaccinated.

First, we focus on conditions when it is possible for ℐ_**V**→_ to become the majority of incidence and find that this is influenced by both assortivity with respect to vaccination status and vaccination coverage. We find that the time until ℐ_**V**→_ is the majority of incidence per day, called *T*_50%_, depends on the assortivity (*ϵ*). In Figure 2A, we simulate Model **G** using parameters from Table S1 in the Supplementary Materials with *ϕ* = 85%, and *ξ* = 30% for various values of assortivity and track unvaccinated-source incidence per day (ℐ_**U**→_) and vaccinated-source incidence per day (ℐ_**V**→_). For *ϵ* = 1, or a population experiencing random mixing, *T*_50%_ = 45 days, and as we introduce assortivity by letting *ϵ* = 0.25 this time increases to *T*_50%_ = 73 days (see Figure 2A). ℐn Figure 2B, we generate a contour of *T*_50%_ with respect to assortivity and the proportion of the population vaccinated, and find there is a threshold proportion of the population vaccinated, *ϕ*, where ℐ_**V**→_ will become the majority of total incidence per day, called *ϕ*_50%_. At low assortivity, *ϵ* = 0.9, this threshold is *ϕ*_50%_ = 43%, and when this threshold is met, the median time until ℐ_**V**→_ is the majority of total incidence per day is *T*_50%_ = 142 days. At *ϵ* = 0.5, the vaccination threshold decreases to *ϕ*_50%_ = 35%, and the median *T*_50%_ = 102 days. At high assortivity, *ϵ* = 0.1, the vaccination threshold further decreases to *ϕ*_50%_ = 3%, and the median *T*_50%_ decreases to *T*_50%_ = 74 days. Note, for perfect assortivity, i.e., vaccinated individuals have a 0% chance of interacting with the unvaccinated, there are no infections in the vaccinated population, and therefore *ϕ*_50%_ does not exist in this case.

**Figure 2:**
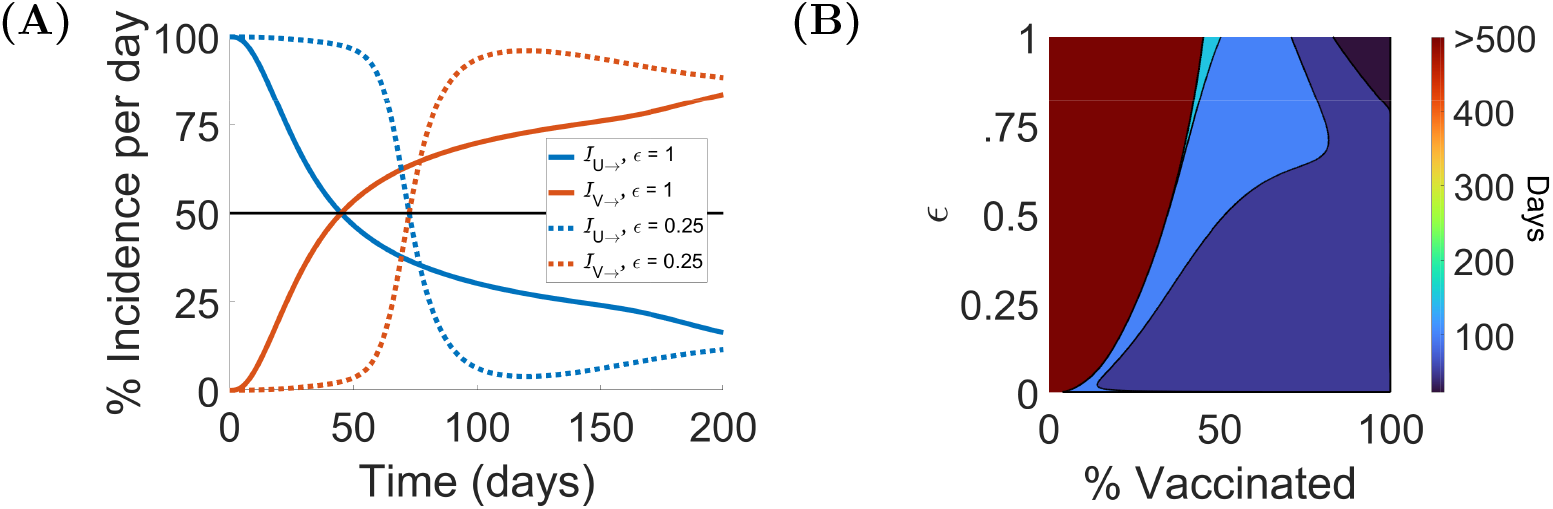
**A. The time until the majority incidence per day is vaccinated-sourced depends on the assortivity.** Percent of incidence from unvaccinated infected individuals, ℐ_**U**→_ (blue), or from vaccinated infected individuals, ℐ_**V**→_ (red), over 200 days for assortivity coefficient values, *ϵ* = 1 and 0.25, generated from Model **G** with parameters from Table S1 in the Supplementary Materials. Note *ϵ* = 1 represents random mixing of vaccinated and unvaccinated populations, while *ϵ* = 0 represents no interaction between vaccinated and unvaccinated populations. **B. There is a threshold proportion of the population vaccinated required for the majority of incidence per day to be from vaccinated infected individuals, and this threshold decreases as assortivity increases**. Contour plot of the time until vaccinated-source incidence is the majority of total incidence per day, *T*_50%_, versus the proportion vaccinated and the assortivity coefficient *ϵ* of the population generated from Model **G** with parameters from Table S1 in the Supplementary Materials. Note, we check for *T*_50%_ for up to 500 days. In cases where ℐ_**V**→_ is not the majority of incidence per day after 500 days, we assume *T*_50%_ does not exist.

Next, we investigate the conditions where ℐ_**V**→_ is the majority of incidence per day during ongoing growth in infections. We identify ongoing growth in infections in our simulations of Model **G** by estimating the effective reproductive number, or number of secondary infections from one infectious individual, with the following equation

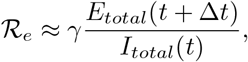

where *γ* is the length of infection. When ℛ_*e*_ *>* 1, there is ongoing growth in infections, or cases are increasing, and when ℛ_*e*_ *<* 1, cases are decreasing. In this study, we focus on conditions where the number of infections in a population are increasing, i.e. ℛ_*e*_ *>* 1. The composition of the vaccinated and unvaccinated infected populations during periods where cases are decreasing but still high may also be relevant for public health interventions, but are not considered for the purposes of this study. For our example, in Figure 1A, there is ongoing growth in infections when *T*_50%_ is reached that continues for 25 days, while ℐ_**V**→_ is the majority of incidence per day. We define three infection spread types in a population of unvaccinated and vaccinated individuals: Type I where ℐ_**V**→_ is never the majority of incidence per day, Type II where ℐ_**V**→_ becomes the majority of incidence per day but ℛ_*e*_ *<* 1 after *T*_50%_, and Type III where ℐ _**V**→_ become the majority of incidence per day and ℛ_*e*_ *>* 1 for some time after *T*_50%_.

We find that the type of infection spread is determined by the assortivity with respect to vaccination status, vaccination coverage, and the length of vaccine-acquired immunity. In Figure 3A, for Model **G** with all other parameters given by Table S1 in the Supplementary Materials, we present the boundaries for Type I, II, and III infection spreads in the *ϵ* − *ϕ* parameter space. Type I infection spread occurs when the proportion of the population vaccinated is below the *ϕ*_50%_ threshold, which is the same boundary found in Figure 2B. A trajectory of incidence per day for a Type I infection spread (*ϵ* = 0.7 and *ϕ* = 10%) is given in Figure 3B, and ℐ_**U**→_ is always the majority of incidence per day. Type II infection spread occurs when *ϕ > ϕ*_50%_ and either assortivity is low or the proportion of vaccinated individuals is moderate. A trajectory of incidence per day for a Type II infection spread (*ϵ* = 0.7 and *ϕ* = 50%) is given in Figure 3C. In this case, ℐ_**V**→_ becomes the majority of incidence per day on *T*_50_ = 121 days, but ℛ_*e*_ had already fallen below one 37 days prior. Type III infection spread occurs when both assortivity and the proportion vaccinated are sufficiently high. A trajectory of new incidence per day for a Type II infection spread (*ϵ* = 0.7 and *ϕ* = 85%) is given in Figure 3D. In this case, ℐ_**V**→_ become the majority of incidence per day on *T*_50_ = 96 days and ℛ_*e*_ *>* 1 for 25 days after. As expected, the peak in incidence per day is decreased and delayed as the proportion vaccinated increases with a value of 3,465 people per day per 100, 000 on day 58 in the Type I example, 1,890 people per day per 100, 000 on day 76 in the Type II example, and 1,139 people per day per 100, 000 on day 114 in the Type III example. In Figure 4, we observed that when the proportion of the population vaccinated is high, as the length of vaccine-acquired immunity is decreased, i.e., faster waning, the assortivity required for Type II rather than a Type III infection spread decreases.

**Figure 3:**
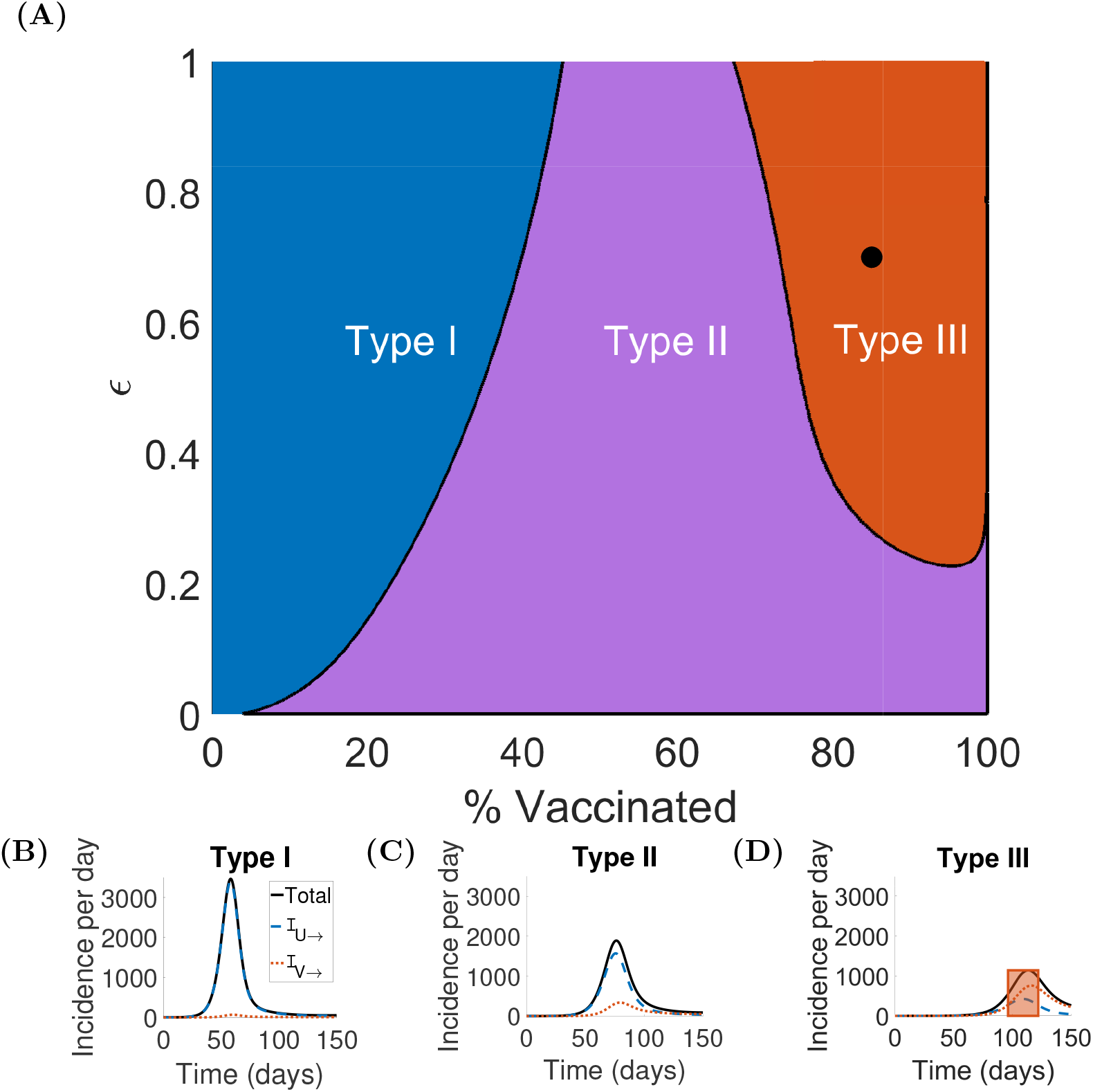
Boundaries defining conditions on the proportion of the population vaccinated and the assortivity that ensure when the majority of incidence will be ℐ_V→_ (Type II and III infection spread) and when the majority of incidence will be ℐ_V→_ during ongoing growth in infections (Type III infection spread). (**A**.) Boundaries defining Types I, II, and III infection spreads defined in the assortivity (*ϵ*) and proportion vaccinated (*ϕ*) parameter space for *ξ* = 0.3 generated from Model **G** with parameters from Table S1 in the Supplementary Materials. The black dot indicates the parameter values used to generate Figure 1A. Sample Type I (**B**.) [*ϵ* = 0.7, *ϕ* = 0.1], Type II (**C**.)[*ϵ* = 0.7, *ϕ* = 0.5], and Type III (D.) [*ϵ* = 0.7, *ϕ* = 0.85] trajectories of total (black) incidence per day, ℐ_**U**→_ (blue), and ℐ_**V**→_ (red). The shaded orange region indicates that ℐ_**V**→_ is the majority of incidence and ℛ_*e*_ *>* 1.

**Figure 4:**
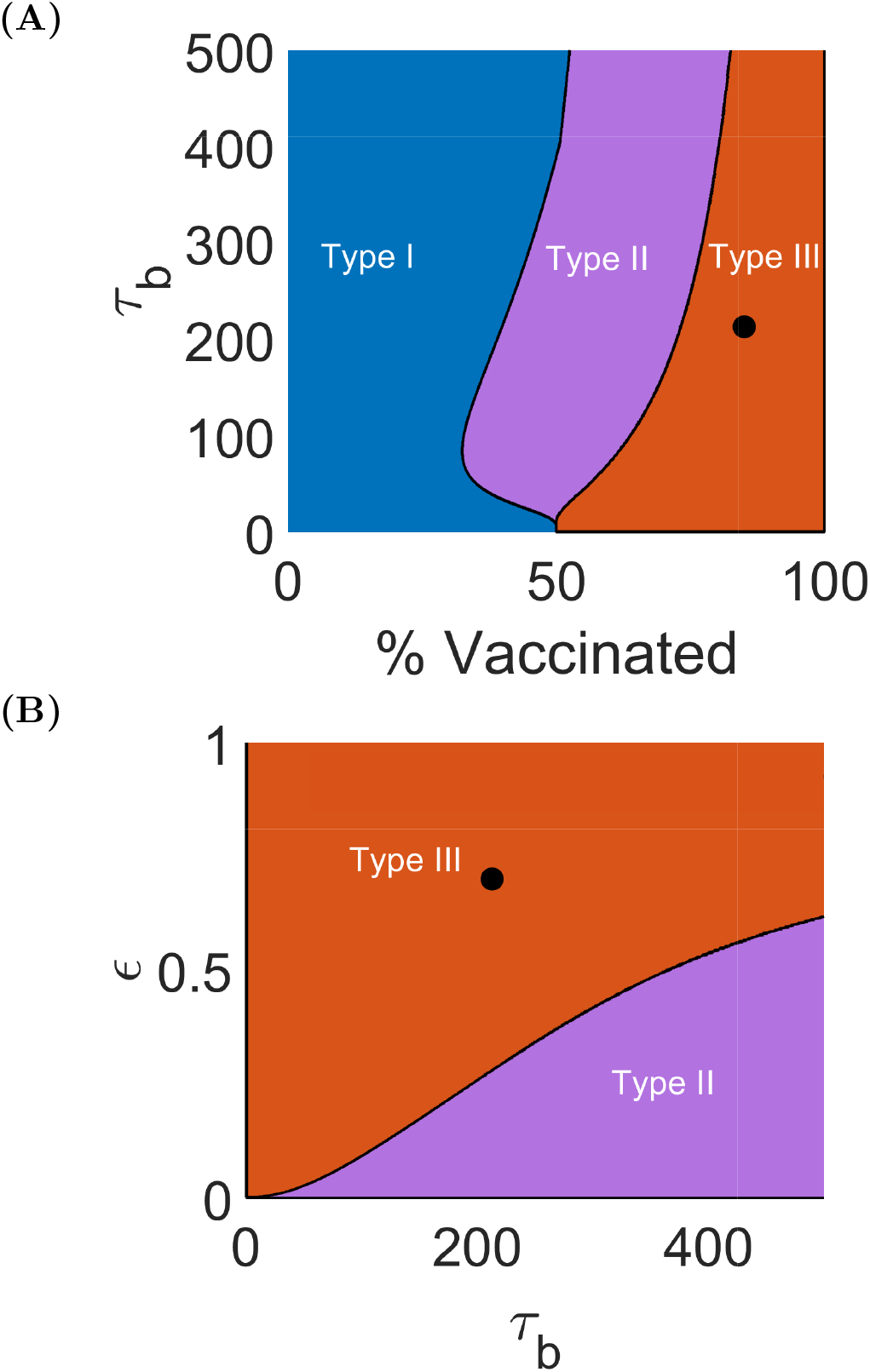
As the length of vaccine-acquired immunity decreases, Type III infection spread can occur at decreasing levels of vaccination coverage and assortivity. (**A**.) Boundaries defining Types I, II, and III infection spread defined in the length of vaccine acquired immunity (*τ*_*b*_) and proportion vaccinated (*ϕ*) parameter space for [*ξ* = 0.3, *ϵ* = 0.7] generated from Model **G** with parameters from Table S1 in the Supplementary Materials. The black dot indicates the parameter values used to generate Figure 1A. (**B**.) Boundaries defining Types I, II, and III infection spread, which are determined by the length of vaccine-acquired immunity (*τ*_*b*_) and assortivity (*ϵ*) parameter space for [*ξ* = 0.3, *ϕ* = 0.85] and parameters given in Table S1 in the Supplementary Materials. Generated from Model **G** with parameters from Table S1 in the Supplementary Materials. The black dot indicates the parameter values used to generate Figure 1A.

We study the impact of masking on incidence per day during Type III infection spread and find, in this case, mask implementation in the vaccinated population is critical to reduce incidence. We simulate Model **G** with parameters from Table S1 in the Supplementary Materials and assume masking implementation reduces transmissions by 50% when both individuals involved in an exposure are masked or by 25% when one individual involved in an exposure is masked. We additionally assume, if masking is implemented, masks are worn by the entirety of the subpopulation for the entire time period considered. Unsurprisingly, we observed the largest increase in delay of the total peak of incidence per day when both unvaccinated and vaccinated populations implement masking, with 72 less incidence per 100, 000 and a delay in peak by 250 days (see Figure 5D), compared to the case where neither population implements masking (see Figure 5A). Under these model conditions, if masking is limited to only one population, it is better to implement masking in the vaccinated population compared to the unvaccinated population. There is a reduction in the peak of incidence per day by 470 people per 100, 000 when masking is implemented in only the vaccinated population (see Figure 5C), while the peak of incidence per day increases by 413 people per 100, 000 if masking is implemented in only the unvaccinated population (see Figure 5B), compared to the case where neither population implements masking. However, the peak in incidence per day is delayed by 50 days when masking is implemented in only the unvaccinated population, but is only delayed by 15 days when masking is implemented in only the vaccinated population (see Figures 5A and B).

**Figure 5:**
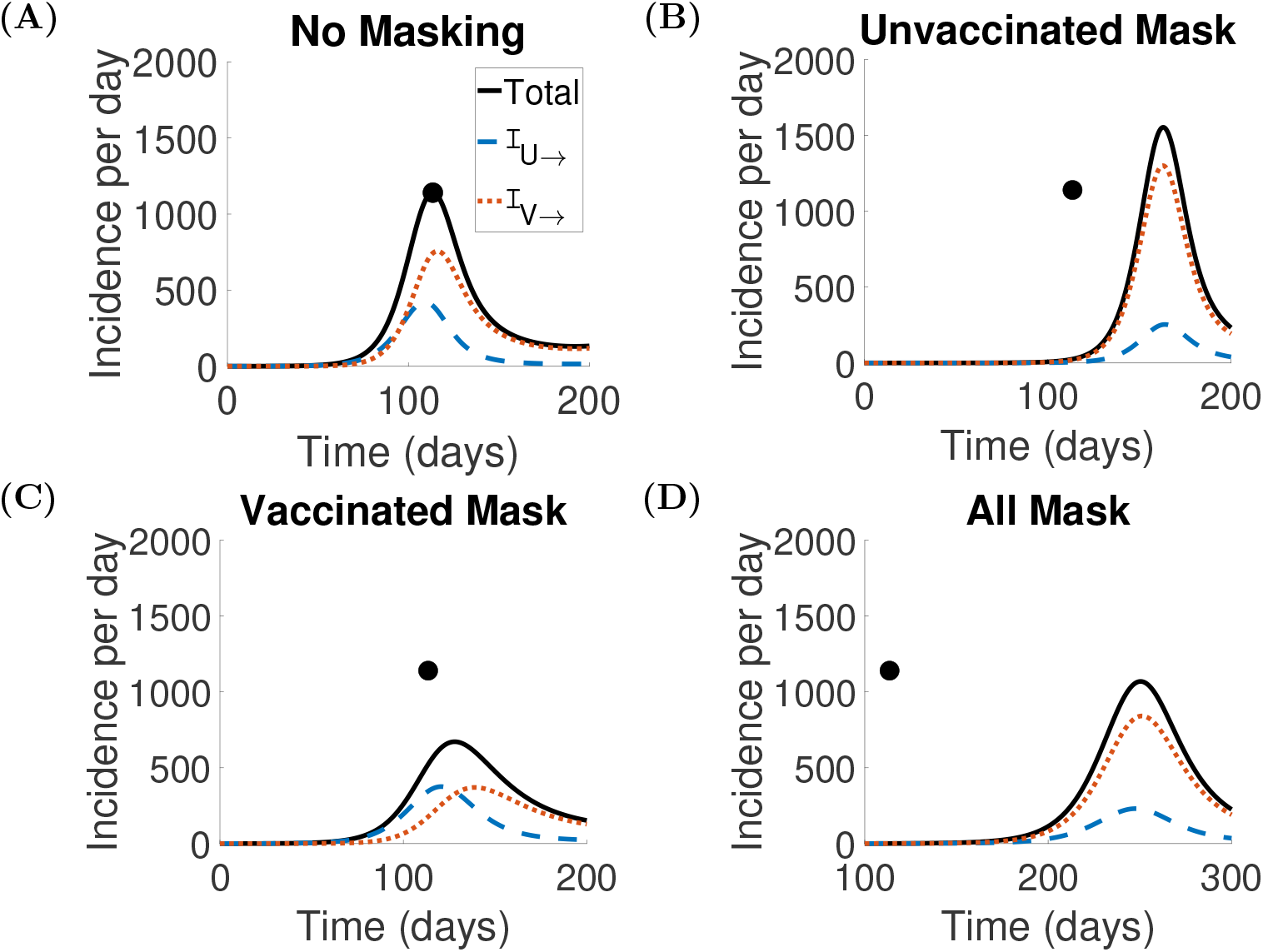
During Type III infection spread, total incidence per day is reduced in cases where the vaccinated population masks, but increased in the case that only the unvaccinated population masks. Total incidence per day per 100,000 (black) over 200 days generated from Model **G** with *ϵ* = 0.7, *ϕ* = 0.85, *ξ* = 0.3 (Type III infection spread), and parameters from Table S1 in the Supplementary Materials. We assume masking reduces transmissions by 50% when both individuals involved in an exposure are masked or by 25% when one individual involved in an exposure is masked. Incidence is classified as arising from interactions with unvaccinated infected individuals, ℐ_**U**→_ (blue), or vaccinated infected individuals, ℐ_**V**→_ (red). **A**. No masking implemented. **B**. Masking is implemented in the unvaccinated population only. **C**. Masking is implemented in the vaccinated population only. **D**. Masking is implemented in both unvaccinated and vaccinated populations. The black circle indicates the peak total incidence per day in the case of no masking.

Moreover, we investigate the impact of masking for examples of both Type I and Type II infection spread and find that the prioritization of which sub-population should mask depends on infection spread type. The peak incidence per day for cases of no masking, unvaccinated only masking, vaccinated only masking, and whole population masking for example choices of Type I, Type II, and Type III infection spread are summarized in Table 2. We observed that masking in the unvaccinated population results in a larger reduction in peak incidence per day (1,791 people per 100,000 vs 42 people per 100,000) than masking in the vaccinated population during a Type I infection spread. During Type II infection spread, masking in only one sub-population results in similar reductions in peak incidence per day, 384 people per 100,000 for unvaccinated masking and 215 people per 100,000 for vaccinated masking. However, only unvaccinated masking contributes to a substantial delay in peak incidence by 65 days.

**Table 2:**
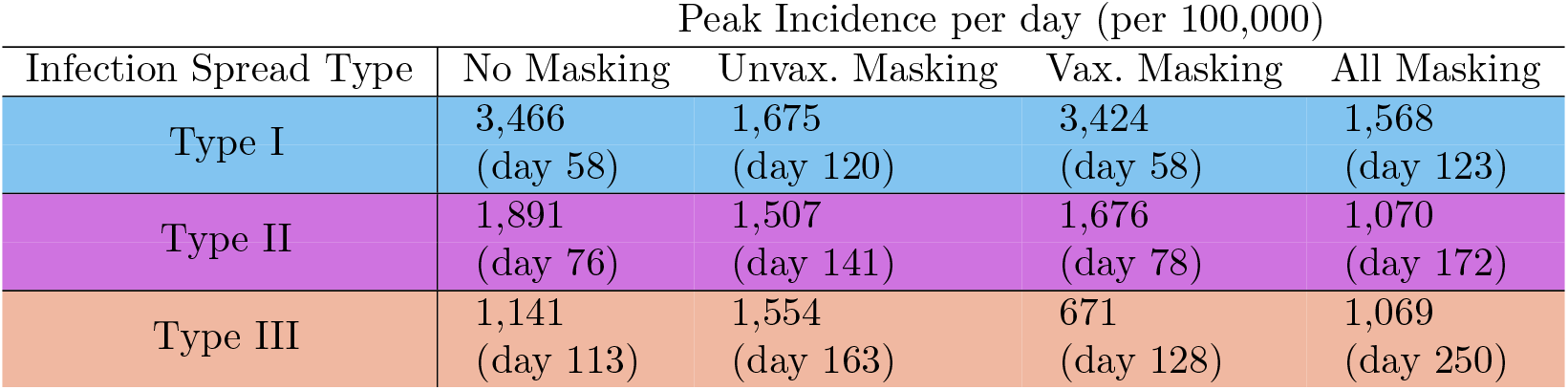
The impact of masking in unvaccinated and vaccinated sub-populations on peak incidence per day changes based on the infection spread type. Maximum incidence per day generated from Model **G** with *ϵ* = 0.7, *ϕ* = 0.1, *ξ* = 0.3 (Type I infection spread), *ϵ* = 0.7, *ϕ* = 0.5, *ξ* = 0.3 (Type II infection spread), and *ϵ* = 0.7, *ϕ* = 0.85, *ξ* = 0.3 (Type III infection spread), and parameters from Table S1 in the Supplementary Materials. We assume masking reduces transmissions by 50% when both individuals involved in an exposure are masked or by 25% when one individual involved in an exposure is masked.

### 2.2 Colorado COVID-19 Transmission Example

Using mathematical modeling, we investigate the contribution of ℐ_**V**→_ to total incidence during the COVID-19 pandemic in Colorado from 2020-2022. Analysis of the model suggests that there were periods during which infected vaccinated individuals contributed to more than 50% of new incidence. In Figure 6A, we fit Model **C** to Colorado COVID-19 hospitalization data collected from March 2020 through December 2022 for assortivity values *ϵ* = [0, 0.05, 0.1, …, 1]. Note assortivity *ϵ* = 0 represents no interaction between unvaccinated and vaccinated subpopulations, while *ϵ* = 1 represents random mixing of unvaccinated and vaccinated individuals. Note that daily hospitalization counts were available through March 2022, after which, weekly counts are used for model parameterization. Biweekly estimates of the transmission rate *β* through March 2022 and monthly estimates of *β* from April 2022 through December 2022 are given in Supplementary Materials, and BIC values ranged between 832 and 898 for models with varying assortivity *ϵ* = [0, 0.05, 0.1, …, 1]. In Figure 6B, we track the incidence per day broken down by ℐ_**U**→_ and ℐ_**V**→_ for the random mixing (*ϵ* = 1) case. Note that vaccine roll-out in Colorado began December 14^*th*^ 2020. As expected from our general transmission model, *T*_50%_ is smallest when we assume the population mixing between unvaccinated and vaccinated individuals is random, *ϵ* = 1, and the earliest vaccine-sourced incidence made up more than 50% of total incidence per day was March 10^*th*^ 2022, 15 months after the start of vaccination. We also find the earliest day for Type III infection spread to have occurred was March 23^*rd*^ 2022. Type III infection spread events are denoted (for the *ϵ* = 1 case) with orange rectangles in Figure 6. There are two time periods from March 23^*rd*^ 2022-June 17^*th*^ 2022 and from September 18^*th*^ 2022-November 7^*th*^ 2022 where growth in infections occurred while ℐ_**V**→_ was the majority of incidence. Also, as expected, in the case that there is no mixing between unvaccinated and vaccinated populations, *ϵ* = 0, there are never infections in the vaccinated population.

**Figure 6:**
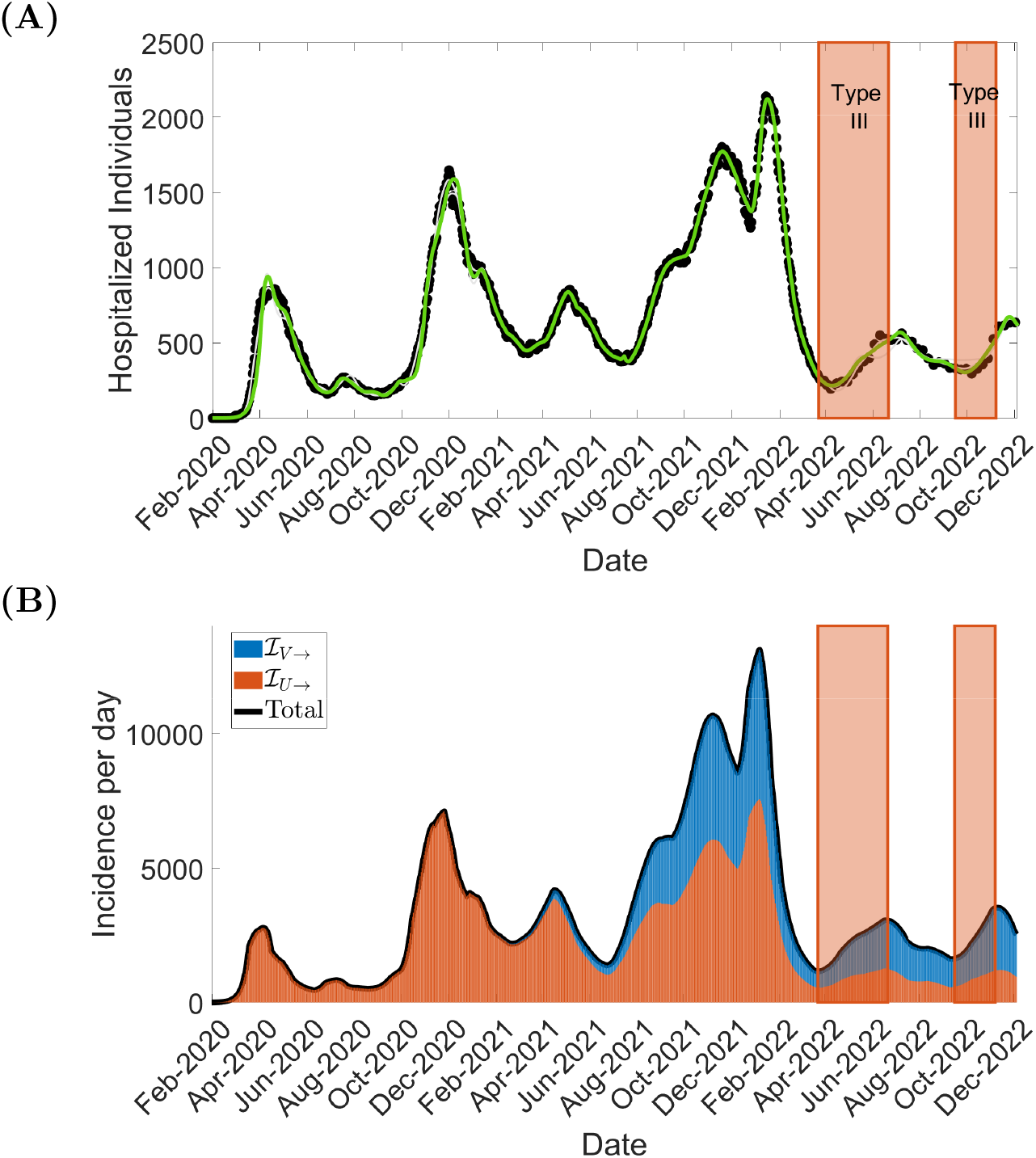
**A. Model C is able to represent Colorado COVID-19 hospitalizations accurately.** COVID-19 hospitalizations in Colorado (black diamond) versus Model B trajectory for *ϵ* = 1 (green line) and *ϵ* ∈ [0, 1] (grey lines) from January 2020 to December 2022. Vaccination in Colorado begins on December 14^th^ 2020. The orange box indicates the model predicted time when ℐ_**V**→_ are the majority of incidence and ℛ_*e*_ *>* 1 for *ϵ* = 1.**B. The mathematical model assuming random mixing predicts the occurrence of Type III infection spread from March** 23^*rd*^ **2022-June** 17^*th*^ **2022 and from September** 18^*th*^ **2022-November** 7^*th*^ **2022**. The incidence per day from unvaccinated, ℐ_**V**→_ (blue), and vaccinated, ℐ_**V**→_ (red), sources between January 2020 and December 2022. Type III infection spread is denoted (for the *ϵ* = 1 case) with orange rectangles. Also, as expected, in the case that there is no mixing between unvaccinated and vaccinated populations, *ϵ* = 0, there are never infections in the vaccinated population.

## 3 Discussion

Using a general compartmental model, we 1) mathematically described three categories of infection spread (Type I, II, III) for a population with waning vaccine-acquired immunity and variable mixing between unvaccinated and vaccinated subpopulations, 2) investigated the impact of additional non-pharmaceutical interventions under these different infection spread conditions, and 3) validated these mathematical predictions by modeling the COVID-19 pandemic in Colorado from 2020-2022.

Using Model **G** (the general model), we demonstrate that when an imperfect vaccine (that does not yield permanent immunity) is introduced and the vaccinated population is sufficiently large, the majority of incidence per day will (for some duration) originate from vaccinated infected individuals. This result is consistent with the work of *Bubar et al*. that found for COVID-19 using compartmental modeling that there is a threshold of vaccination coverage where the majority of infections will be in the vaccinated population [32]. Furthermore, we find this threshold is relatively low (less than 45%) across various mixing levels between vaccinated and unvaccinated populations. We note that this threshold is dependent on the length of vaccine-acquired immunity, as demonstrated in Figure 4A. While outside the scope of our model assumptions, we conjecture that it would be unlikely for Type III infection spread to occur in the case that vaccine-acquired immunity endures much longer than infection acquired immunity.

More significantly, we showed that there are circumstances where ℐ_**V**→_ over-takes ℐ_**U**→_ during an ongoing period of growth in infections. We define this scenario as a Type III infection spread, including conditions on the assortivity, proportion vaccinated, and length of vaccine-acquired immunity are depicted in Figure 3. Distinguishing between Type I, II, and III infection spread is important because interventions in the unvaccinated or vaccinated population will impact the ongoing growth in infections differently based on infection spread type. As demonstrated in Table 2, to best reduce (or delay) peak incidence in Type I infection spread, it is most important for unvaccinated individuals to mask, while to best reduce peak incidence in Type III infection spread, it is most important for the vaccinated to mask. While for all infection spread types, masking in the whole population results in the largest reduction in peak incidence, it is the critical intervention for making impactful reductions in incidence in Type II infection spread. Recall, as demonstrated in Figure 1B and the example cases presented in Figure 3B-D, that increases in vaccination coverage are always beneficial in reducing total cases and delaying peak incidence per day. Therefore, the classification of infection spread into Type I, II, and III provides guidance on supplementary interventions that can provide additional control of disease spread.

Other supplementary interventions that we can study through the infection spread type classification are policies restricting access to spaces based on vaccination status. In our model, assortivity, or sub-population mixing, can also be altered by such policies. Starting in Type I or Type II infection spread conditions, these policies would not impact the infection dynamics, meaning ℐ_**V**→_ will never be the majority of incidence during a period of growth in infections. However, starting in Type III infection spread conditions, these isolation policies could result in a Type II infection spread instead. Concerning the Colorado COVID-19 model (**C**), we were able to verify our results from Model **G** by predicting two periods during the pandemic where growth in infections could be classified as Type III (dependent on the assumption of mixing between unvaccinated and vaccinated populations), meaning the majority of incidence per day was vaccinated-source incidence during active growth in infections. We fit the Colorado-specific Model **C** to hospitalization data from January 2020 to December 2022, and then simulated Model **C** for various mixing between vaccinated and unvaccinated populations. The proportion of the population vaccinated in Colorado reached over 40% by May 2021. Based on our work with Model **G**, we would expect that there will be a point after May 2021 where the majority of incidence per day in Colorado would be vaccinated-source incidence, and indeed, this phenomenon is observed using Model **C**. ℐ_**V**→_ were the majority of incidence per day starting between March 10^th^ 2022 and May 4^th^ 2022. We have given one example where the predictions from Model **G** hold for a real-life disease spread of a respiratory infection. We anticipate these results to hold for a broad variety of respiratory infections when vaccination is widely available, but vaccine-acquired immunity is waning.

Our study has several limitations. First and foremost, it is important to note that the conclusions in this study about incidence involving vaccinated individuals are predicted from mathematical models. In particular, the predictions from Model **C** for transmission during the COVID-19 pandemic directly arise from analysis of the model. Unfortunately, validation of the predictions could only have been confirmed through contact tracing during that time. While there have been documented cases of transmission to and transmission by vaccinated individuals during the COVID-19 pandemic [33, 34], there has not been a widespread study to confirm the frequency of these occurrences. Indeed, it would be disingenuous to conclude that the pandemic could be controlled via altering a single aspect of the behaviors of vaccinated individuals, e.g., mandating masking for vaccinated individuals.

Secondly, we assume the average lengths of protection from both vaccine and infection-acquired immunity are the same over a population; however, there is likely variability in this length of protection, particularly for vulnerable populations. Additionally, there may be behavioral differences between the unvaccinated and vaccinated populations that impact their respective roles in transmission during periods of growth in infections. In particular, one might anticipate a difference in compliance with isolation or masking recommendations between these two groups. More work is needed to understand the impact of these potential behavioral differences.

Additionally, this study suggests that reducing transmission in the vaccinated sub-population is important in reducing peak incidence in Type III infection spread. We investigated one method of reducing transmission by simulating the effects of masking in the vaccinated population. In this study, we assume a 50% reduction in transmission during implementation of masking. Masks have been shown to be effective at blocking transmission of SARS-CoV-2 [35, 36], however, our work did not consider population behavior. It is unlikely that a whole population would wear, or properly wear, masks as shown for the general model. We find in Table S2 (see Supplementary Materials), for cases where masks cause less reduction in transmission, that infection spread type still influences whether to require masks in a sub-population or the whole population, but that the reductions in incidence and delay in peak incidence gained from masking are less substantial. Another method of transmission reduction is the vaccine itself. A density-dependent relationship between SARS-CoV-2 viral load and infectiousness of a host individual has been suggested by [37, 38], and COVID-19 vaccinations have been observed, dependent on the strain, to reduce the time viral load is elevated, although maximum viral loads may be similar between unvaccinated and vaccinated [39–41]. Further, recently there has been progress in developing nasal vaccines, which may prove more effective at preventing transmission [42, 43]. In fact, for vaccines that are highly effective at blocking transmission, we would only expect Type III infection spread at both high vaccination rates and high population mixing, as demonstrated in Figure S5 in the Supplementary Materials.

Finally, uncertainty quantification is critical to using model predictions for infectious disease decision-making [44–47]. An important factor affecting uncertainty when using compartmental models includes parameter identifiably, which can be heavily influenced by noise and data availability [48–50]. More careful consideration of uncertainty quantification and parameter identifiability would be needed to use our modeling framework for future predictions of periods where vaccinated-involved incidence drives growth in infections.

In conclusion, we defined three infection spread types: where ℐ_**V**→_ is never the majority of incidence (Type I), where ℐ_**V**→_ becomes the majority of incidence but not during a period of growth in infections (Type II), and where ℐ_**V**→_ is the majority of incidence during a period of growth in infections (Type III). We determined parameter spaces where these three infection spread types will occur in terms of assortivity, vaccination coverage, and length of vaccine-acquired immunity, found that infection spread type impacts the effectiveness of population and sub-population level masking implementation, and demonstrated, even in circumstances of high vaccination, that requiring only the unvaccinated to mask can increase the total exposures per day. Finally, we verified these results by showing that ℐ_**V**→_ were the majority of new COVID-19 incidence between March 23^*rd*^ 2022-June 17^*th*^ 2022 and from September 18^*th*^ 2022-November 7^*th*^ 2022 in Colorado. Ultimately, our work suggests that it is necessary to consider waning vaccinated immunity when implementing supplementary interventions, such as masking, to prevent new infections.

## 4 Methods

In this section, we will describe mathematical models to investigate the impact of waning vaccine-induced immunity on the daily incidence rate. We will first describe a general (idealized) model, followed by a full age-structured model of the COVID-19 pandemic in Colorado during 2020-2022. We chose this period as it coincides with when the first wave of mRNA-based vaccines became available. We note that for the convenience of the reader, Tables S1 and S2 (see Supplementary Materials) collect all the relevant variables and parameters.

### 4.1 General Transmission Model

First, we model the spread of a human-to-human transmitted disease using a general SEIR-type compartmental model. As depicted in the diagram in Figure 7, the total population *N* is composed of two sub-populations *N* ^U^ and *N* ^V^, for which we denote the unvaccinated with a superscript U and the vaccinated with a superscript V. We begin with a population of vaccinated individuals *V* who can return to the susceptible population *S*^V^. Vaccine effectiveness at blocking transmission is assumed to be 100%. From there, vaccinated *S*^V^ and unvaccinated *S*^U^ susceptibles pass through exposed, infectious, and recovered categories before becoming susceptible again. To isolate the impact of the U and V sub-populations on incidence, in this model, we do not allow for unvaccinated individuals to become vaccinated. Naturally, in the Colorado COVID-19 model in Section 4.2, we do include terms describing ongoing vaccinations.

**Figure 7:**
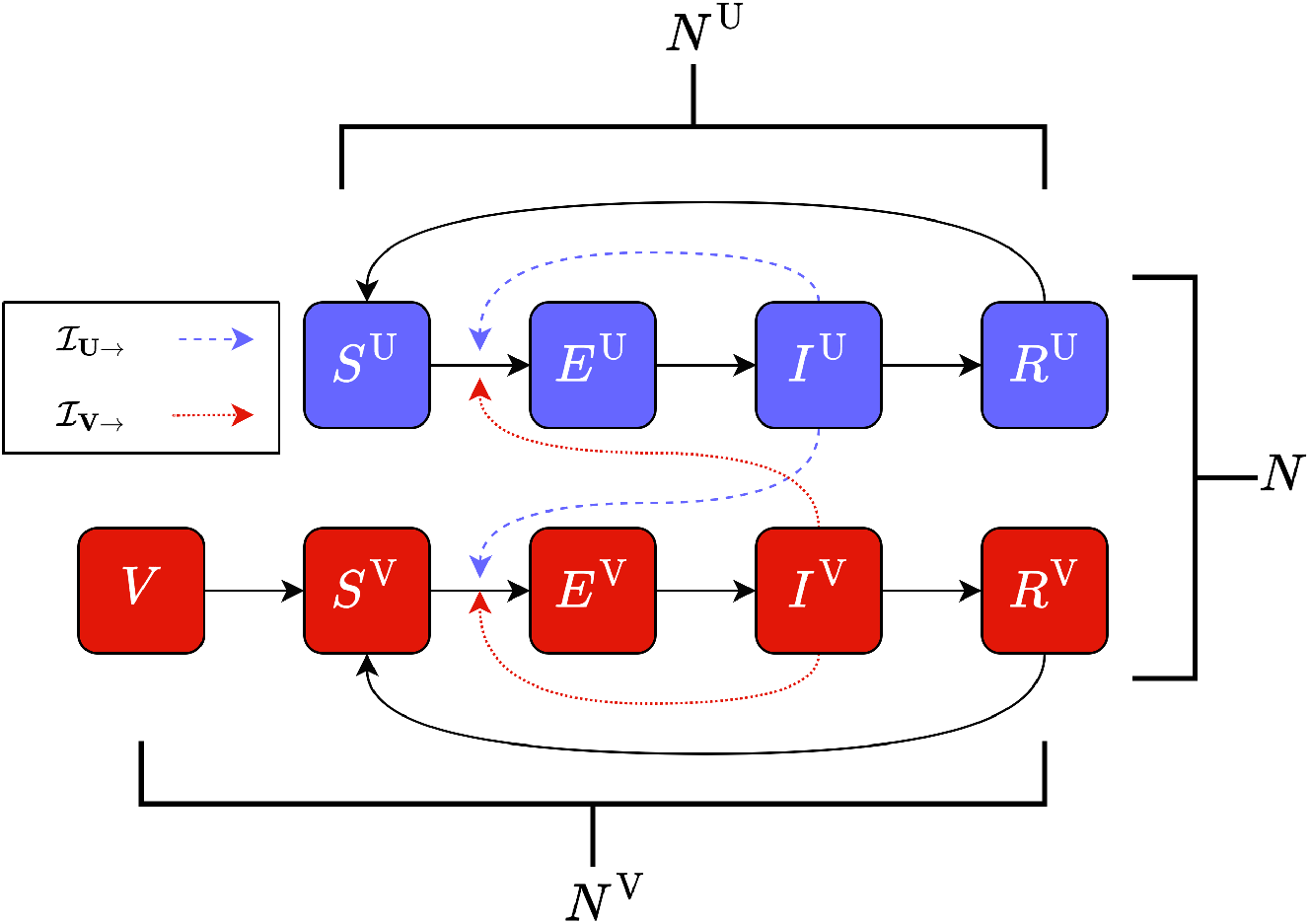
Schematic diagram of Model **G**. Dashed lines represent new transmissions that arise from an unvaccinated source, i.e. ℐ_**U**→**U**_ and ℐ_**U**→**V**_ (blue dash), and those that arise from a vaccinated source, i.e. ℐ_**V**→**U**_ and ℐ_**V**→**V**_ (red dot).

The general transmission model, Model **G**, is given by Eq. (S1), with parameter values given in Table S1, in the Supplementary Materials.

We include terms describing preferential mixing and follow the method in [51] to construct the matrices describing mixing between unvaccinated and vaccinated sub-populations. The proportion of contacts an individual experiences that are strictly within their sub-population is given by (1 −*ϵ*), and the proportion of random contacts is given by *ϵ*. We call *ϵ* the *assortivity constant*, and when *ϵ* = 1 there is only random mixing, while *ϵ* = 0 means that the subpopulations do not interact.

Aside from the modification to account for mixing, the terms in (**??**) are all standard SEIR-type model terms. For *i* ∈ {U, V}, Susceptibles become Exposed *E*^*i*^ at rate *β/N* ^*i*^ per day for contacts with the Infectious *I*^*i*^ and rate *β/N* per day for random contacts with all Infectious. Individuals remain Exposed (on average) for *α* days before becoming Infectious and Recover (on average), *R*^*i*^, in *γ* days. The Recovered maintain immunity to re-infection for an average of *τ*_*i*_ days. The vaccinated subpopulation starts in an additional Vaccinated compartment, *V*, and has immunity to infection for an average of *τ*_*b*_ days.

The total population and the subpopulations remain constant, meaning, *N* ^U^ = *S*^U^ + *E*^U^ + *I*^U^ + *R*^U^, *N* ^V^ = *V* + *S*^V^ + *E*^V^ + *I*^V^ + *R*^V^, and *N* = *N* ^V^ + *N* ^U^. We assume the following initial conditions, which represent a population with initial infection 16 acquired immunity, 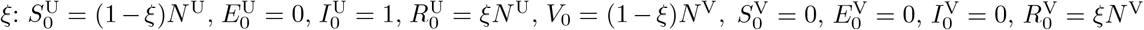

Our main focus in this work is on understanding the relationship between the dynamics of new transmission events and the effects of waning vaccinated immunity. Accordingly, we partition the number of new transmissions per day or total incidence per day (ℐ_**T**_) into four categories based on the vaccination status of the individuals participating in the transmission event. We designate ℐ_**U**→**U**_ as the incidence occurring from two unvaccinated individuals, ℐ_**U**→**V**_ as the incidence occurring from an unvaccinated infected and vaccinated susceptible individual, ℐ_**V**→**U**_ as the incidence occurring from a vaccinated infected and unvaccinated susceptible individual, and ℐ_**V**→**V**_ as the incidence occurring from two vaccinated individuals. The formulae are given as:

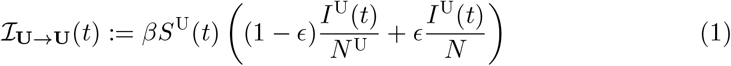

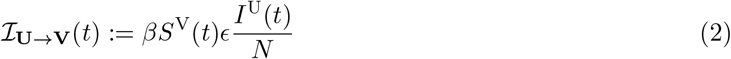

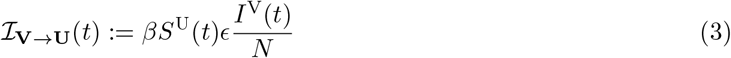

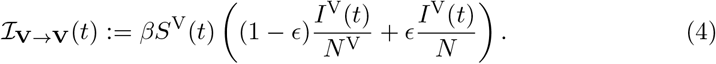

and we note that ℐ_**T**_ = ℐ_**U**→**U**_ + ℐ_**U**→**V**_ + ℐ_**V**→**U**_ + ℐ_**V**→**V**_.

In this work, we partition ℐ_**T**_ by the vaccination status of the infected individual or the source. We let ℐ_**T**_ = ℐ_**U**→_ + ℐ_**V**→_, where ℐ_**U**→_ is the unvaccinated-source incidence and ℐ_**V**→_ is the vaccinated-source incidence, i.e.,

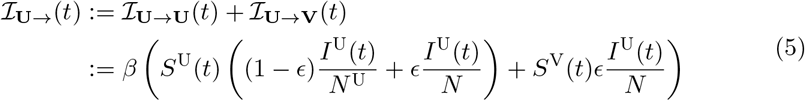

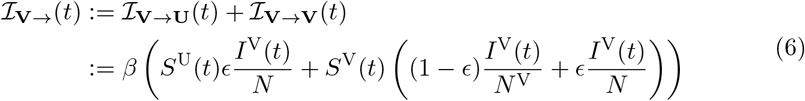

### 4.2 Colorado COVID-19 Transmission Example

We next model COVID-19 spread in Colorado from January 2020 through December 2022 by expanding Model **G**. In the Colorado-specific model, Model **C**, we additionally consider compartments for (where *i* ∈ {U, V}) Asymptomatic *A*^*i*^, Hospitalized *H*^*i*^, Boosted *B*, and Deceased *D* individuals. As depicted in the diagram in Figure 8, individuals may become Asymptomatic or Infected after being Exposed, and Infected individuals may either Recover or become Hospitalized, while Asymptomatics only Recover. Hospitalized individuals either Recover or Die, and both Recovered and Susceptible individuals can become Vaccinated or Boosted. Unlike in the general model presented in 4.1, where vaccination status is determined at day 1 and no new vaccination can occur, this model simulates a roll-out of new vaccinations over time to mirror real-world vaccination campaigns.

**Figure 8:**
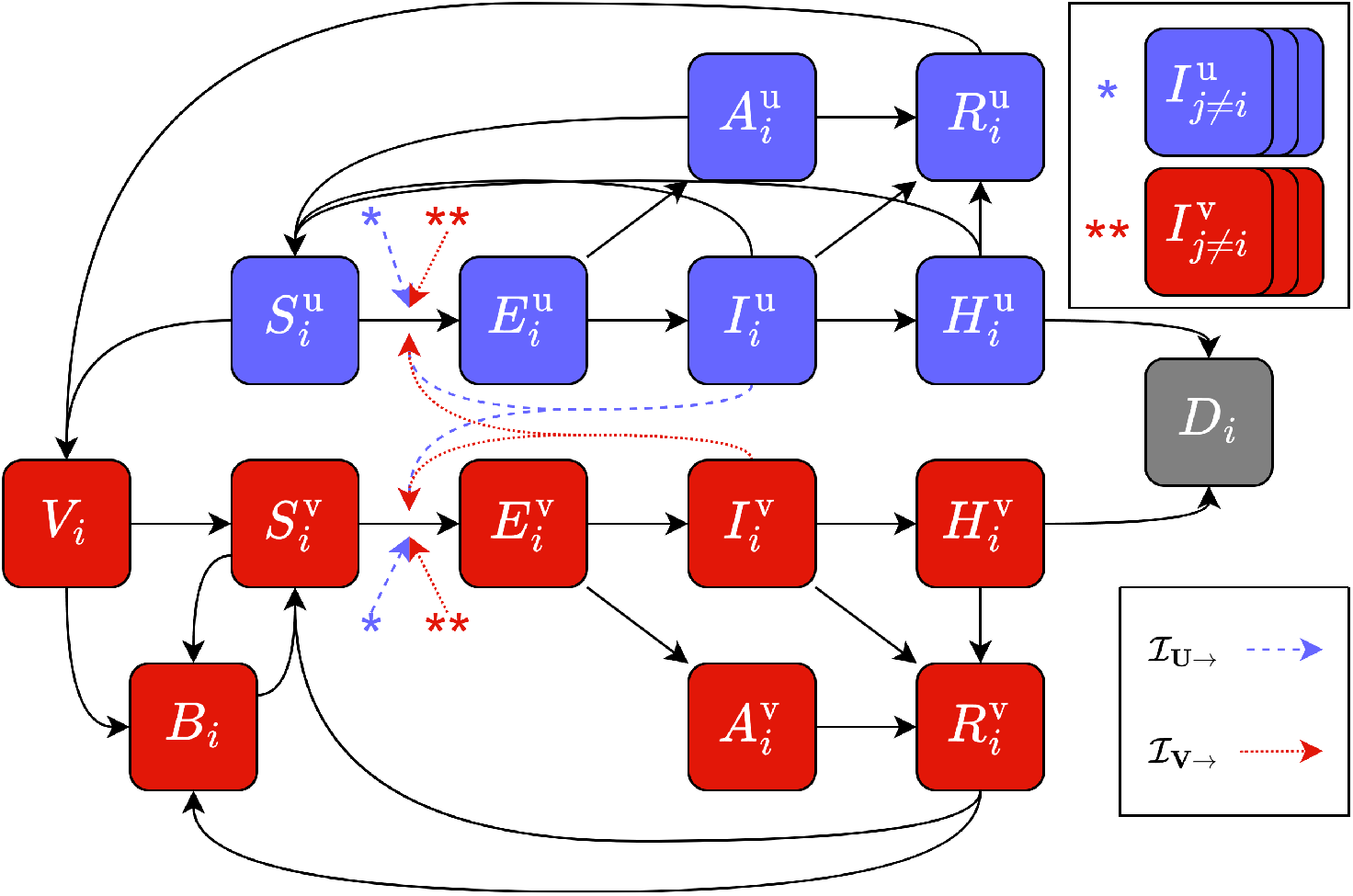
Schematic diagram of Model **C** for a single age class *i*. Dashed lines represent new transmissions that arise from an unvaccinated source, i.e. ℐ_**U**→**U**_ and ℐ_**U**→**V**_ (blue dash), and those that arise from a vaccinated source, i.e. ℐ_**V**→**U**_ and ℐ_**V**→**V**_ (red dot).

The Colorado-specific model, Model **C**, equations are given by Eq. (S2) and parameter values are given in Table S3 in the Supplementary Materials. The formula for ℐ_**U**→**U**_, ℐ_**U**→**V**_, ℐ_**V**→**U**_, and ℐ_**V**→**V**_ for Model **C** are given by Eqs. (S3)-(S6) in the Supplementary Materials.

As in Model **G**, we assume preferential mixing between unvaccinated and vaccinated sub-populations. We further assume symptomatic infectious individuals have a higher transmission rate, *β*_*I*_, than asymptomatic individuals that transmit at rate *β* [52]. Additionally, to represent individuals with compromised immune response, in the unvaccinated subpopulation, small fractions of Asymptomatic individuals, *λ*_*A*_, and of Infected or Hospitalized individuals, *λ*_*I*_, become Susceptible to reinfection without a recovery period. To better represent the population of Colorado we additionally assume the fraction of symptomatic infections, *ρ*, the rate of non-hospitalized deaths, *d*_*nh*_, the rate of hospitalized deaths, *d*_*h*_, the rate of hospitalization, *h*, and the length of hospitalization, *s*, differ based on the following age categories 0 to 19, 20 to 39, 40 to 64, and 65+. Furthermore, we adjust the length of hospitalization and hospitalization rates throughout the time frame of our model to account for both policy changes and the introduction of new SARS-CoV-2 variants in Colorado.

We no longer enforce that the total population or sub-populations remain constant because we allow for both death and movement from the unvaccinated sub-population to the vaccinated sub-population. However, at any given time 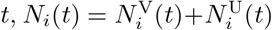 for age classes *i* = {0 to 19, 20 to 39, 40 to 64, and 65+} and 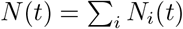

We use Colorado daily hospitalization data collected from March 2020 through April 2022 and weekly hospitalization data collected from April 2022 through December 2022 from the Colorado Department of Public Health and Environment open database [53], and we begin simulations of Model **C** on January 1st, 2020, assuming 1 initially infected individual.

To model COVID-19 spread in Colorado, we estimate the transmission rate, *β*, on bi-weekly intervals by fitting Model **C** to Colorado daily hospitalization data, and we consider all other parameters to be fixed to the values given in Table S3 in the Supplemental Materials. The model is implemented as a simBiology model in MATLAB [54], and we use the sbiofit tool to perform a hybrid optimization to produce these estimates, first applying the global particle swarm optimizer based on [55] as implemented in the `particleswarm` command and then performing a local minimization with `fmincon`.

## Data Availability

All data produced in the present work are contained in the manuscript.

## 5 Acknowledgements

Research reported in this publication was supported in part by the National Institute of General Medical Sciences of the National Institutes of Health under award number R35GM149335. The authors also received support from the Colorado Department of Public Health and Environment. The content is solely the responsibility of the authors and does not necessarily represent the official views of the aforementioned funding organizations.

The authors would also like to thank S. Kissler (University of Colorado) for insightful comments on an early draft of this work.

## 6 Author Contributions

S.A. and D.M.B. conceived the idea. N.H.B., S.A., and D.M.B. designed the study, analyzed the results, and were responsible for the initial drafting of the manuscript. N.H.B. and S.A. developed the code. N.H.B generated the results and figures. D.M.B. and E.C. supervised the project. j.a. and A.G.B contributed to model development. All of the authors contributed to the writing and editing of the manuscript.

## 7 Code Availability

The code for this paper will be provided on Github at https://github.com/MathBioCU/VaccinationImmunityModeling.

