## Supplemental Materials for "Waning Immunity and Partial Vaccination Coverage Lead to Transitions in the Source of Daily Incidence"

### S1: General Transmission Model

The general transmistion model, Model **G**, is given by the equation below,

$$
\begin{aligned}
\frac{dS^{\mathrm{U}}}{dt} &= -\beta S^{\mathrm{U}}\left[(1-\epsilon)\left(\frac{I^{\mathrm{U}}}{N^{\mathrm{U}}}\right)+\epsilon\left(\frac{I^{\mathrm{V}}+I^{\mathrm{U}}}{N}\right)\right]+\frac{1}{\tau_{\mathrm{U}}}R^{\mathrm{U}}\\
\frac{dE^{\mathrm{U}}}{dt} &= \beta S^{\mathrm{U}}\left[(1-\epsilon)\left(\frac{I^{\mathrm{U}}}{N^{\mathrm{U}}}\right)+\epsilon\left(\frac{I^{\mathrm{V}}+I^{\mathrm{U}}}{N}\right)\right]-\frac{1}{\alpha}E^{\mathrm{U}}\\
\frac{dI^{\mathrm{U}}}{dt} &= \frac{1}{\alpha}E^{\mathrm{U}}-\frac{1}{\gamma}I^{\mathrm{U}}\\
\frac{dR^{\mathrm{U}}}{dt} &= \frac{1}{\gamma}I^{\mathrm{U}}-\frac{1}{\tau_{\mathrm{U}}}R^{\mathrm{U}}\\
\frac{dV}{dt} &= -\frac{1}{\tau_b}V\\
\frac{dS^{\mathrm{V}}}{dt} &= -\beta S^{\mathrm{V}}\left[(1-\epsilon)\left(\frac{I^{\mathrm{V}}}{N^{\mathrm{V}}}\right)+\epsilon\left(\frac{I^{\mathrm{V}}+I^{\mathrm{U}}}{N}\right)\right]+\frac{1}{\tau_b}V+\frac{1}{\tau_{\mathrm{V}}}R^{\mathrm{V}}\\
\frac{dE^{\mathrm{V}}}{dt} &= \beta S^{\mathrm{V}}\left[(1-\epsilon)\left(\frac{I^{\mathrm{V}}}{N^{\mathrm{V}}}\right)+\epsilon\left(\frac{I^{\mathrm{V}}+I^{\mathrm{U}}}{N}\right)\right]-\frac{1}{\alpha}E^{\mathrm{V}}\\
\frac{dI^{\mathrm{V}}}{dt} &= \frac{1}{\alpha}E^{\mathrm{V}}-\frac{1}{\gamma}I^{\mathrm{V}}\\
\frac{dR^{\mathrm{V}}}{dt} &= \frac{1}{\gamma}I^{\mathrm{V}}-\frac{1}{\tau_{\mathrm{V}}}R^{\mathrm{V}}
\end{aligned}
\tag{S1}
$$

#### S1.1 Variable Mixing Between Sub-Populations

We consider the total population of size $N$ to be divided into two sub-populations, vaccinated and unvaccinated. While $N$ remains constant for all time, the vaccinated population $N^{\mathrm{V}}$ increases, and the unvaccinated population $N^{\mathrm{U}}$ decreases when new vaccines are administered. We will stick to the relevant two sub-population scenario; however, the generalization to $n$ sub-populations is natural.

The mixing pattern between $n$ sub-populations is expressed by an $n \times n$ contact matrix $\mathcal{M} = (m_{xy})$. Entries $m_{xy}$ give the probability that a member of group $x$ comes in contact with a member of group $y$. We parameterize $(m_{xy})$ by $\epsilon$, the degree of assortativity, such that mixing is random when $\epsilon = 1$, and fully assortative when $\epsilon = 0$. For random mixing, each entry $m_{xy}$ should be the fraction of the population made up by sub-population $y$. When mixing is completely assortative, $\mathcal{M}$ should reduce to the identity matrix with elements defined by $\delta_{xy}$, the Kronecker delta function. Thus, we define elements of $\mathcal{M}$ to be,

$$m_{xy} = (1-\epsilon)\delta_{xy} + \epsilon\left(\frac{N^y}{N}\right),$$

| parameter | description | unit | value |
|---|---|---|---|
| $\beta$ | transmission rate | per day | 0.75 |
| $\alpha$ | incubation period | days | 4 |
| $\gamma$ | length of infection | days | 9 |
| $\tau_{\mathrm{U}}$ | length of protection from infection acquired immunity | days | 365 |
| $\tau_b$ | length of protection from vaccine acquired immunity | days | 213 |
| $\tau_{\mathrm{V}}$ | length of protection from infection acquired immunity after vaccination | days | 365 |
| $N$ | total population | people | 100,000 |
| $\epsilon$ | assortivity between vaccinated and unvaccinated populations | - | $[0, 1]$ |
| $\xi$ | proportion of initial population with infection acquired immunity | - | $[0, 1]$ |
| $\phi$ | proportion of population vaccinated | - | $[0, 1]$ |

**Table S1**: Parameter values used in simulations of Model **G**. For parameters $\epsilon$, $\xi$, and $\phi$ the values listed above represent the lower and upper bounds.

where again, $N^y$ is the size of sub-population $y$ and $N$ is the total population. For the two sub-populations $u$ and $v$ designated by vaccination status, the contact matrix $\mathcal{M}$ becomes,

$$(m_{uv}) = \begin{bmatrix} (1-\epsilon) + \epsilon\left(\frac{N^u}{N}\right) & \epsilon\left(\frac{N^v}{N}\right) \\ \epsilon\left(\frac{N^u}{N}\right) & (1-\epsilon) + \epsilon\left(\frac{N^v}{N}\right) \end{bmatrix}.$$

In a simple SIR or SEIR model with random mixing, the ODE describing the rate of change in the susceptible population is,

$$\frac{dS}{dt} = -\beta S \frac{I}{N},$$

with transmission rate $\beta$. We include non-random mixing by amending the fraction of infectious contacts. Instead of $\frac{I}{N}$, the non-random fraction of infectious contacts is the sum of the probability of contact with each group, multiplied by the fraction of that group that is infectious.

For our two sub-population model, the expression for the unvaccinated susceptible population becomes,

$$\frac{dS^{\mathrm{U}}}{dt} = -\beta S^{\mathrm{U}} \left[\left((1-\epsilon) + \epsilon\left(\frac{N^{\mathrm{U}}}{N}\right)\right)\frac{I^{\mathrm{U}}}{N^{\mathrm{U}}} + \epsilon\left(\frac{N^{\mathrm{V}}}{N}\right)\frac{I^{\mathrm{V}}}{N^{\mathrm{V}}}\right],$$

and when reduced to,

$$\frac{dS^{\mathrm{U}}}{dt} = -\beta S^{\mathrm{U}} \left[(1-\epsilon)\frac{I^{\mathrm{U}}}{N^{\mathrm{U}}} + \epsilon\left(\frac{I^{\mathrm{U}} + I^{\mathrm{V}}}{N}\right)\right].$$

##### S1.2 Effect of assortivity

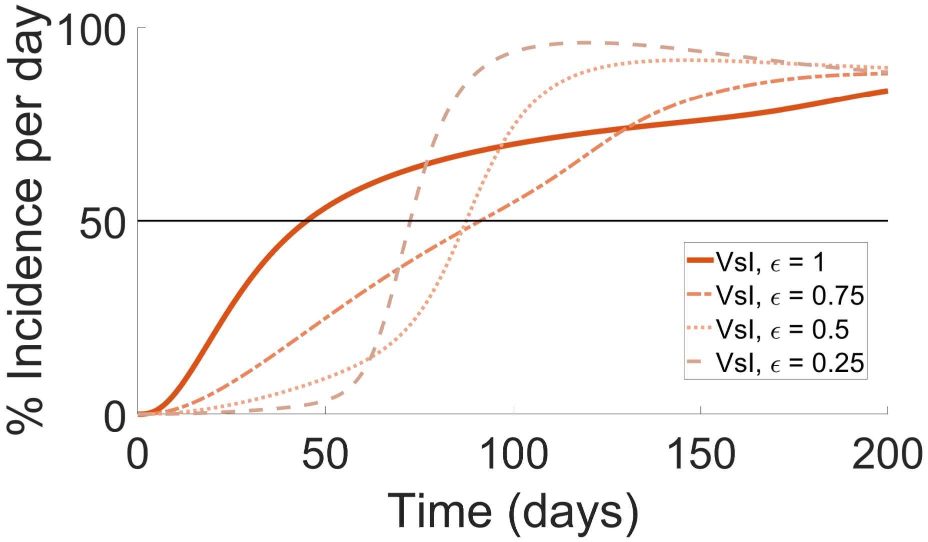


**Figure S1: As assortivity increases ($\epsilon$ decreases), the proportion of incidence per day from vaccinated infected individuals over time becomes more sigmoidal.**

##### S1.3 Effect of initial proportion of naturally acquired immunity

Unlike the relationship between assortivity and $\phi_{50\%}$, as the proportion of the population with infection acquired immunity, $\xi$, increases the threshold proportion of the population vaccinated where $\mathcal{I}_{\mathbf{U}\to\mathbf{V}}+\mathcal{I}_{\mathbf{V}\to\mathbf{U}}+\mathbf{VI}$ becomes the majority of total incidence strictly increases (see Figure S2A). At high initial natural immunity, $\xi = 90\%$, the vaccination threshold is $\phi_{50\%} = 36.5\%$, and the median time until $\mathcal{I}_{\mathbf{U}\to\mathbf{V}}+\mathcal{I}_{\mathbf{V}\to\mathbf{U}}+\mathbf{VI}$ is the majority of total incidence is $T_{50\%} = 93$ days (see Figure S2B). At $\xi = 50\%$, the threshold decreases to $\phi_{50\%} = 29.5\%$, and the median $T_{50\%}$ decreases to $T_{50\%} = 77$ days (see Figure S2C). Finally, at low initial natural immunity, $\xi = 10\%$, the threshold decreases to $\phi_{50\%} = 22.5\%$, and the median $T_{50\%}$ decreases to $T_{50\%} = 66$ days (see Figure S2D).

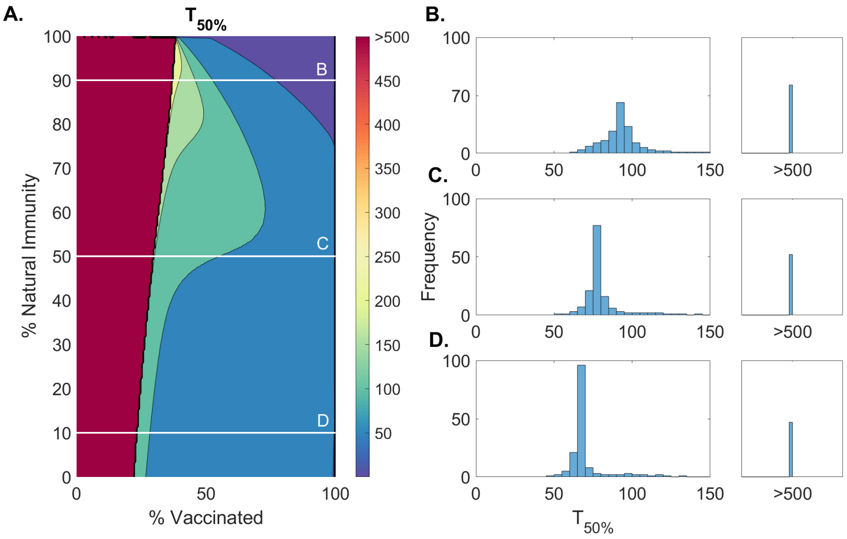


**Figure S2**: **A.** Contour plot of the time until vaccinated-sourced incidence is the majority of total incidence, $T_{50\%}$, versus the proportion vaccinated and the proportion of the population with infection acquired immunity generated from Model **G**. **B.** Histogram of $T_{50\%}$ for simulations with various proportions vaccinated and 90% infection acquired immunity represented by the white line in (A.). **C.** Histogram of $T_{50\%}$ for simulations with various proportions vaccinated and 50% infection acquired immunity represented by the white line in (A.). **D.** Histogram of $T_{50\%}$ for simulations with various proportions vaccinated and 10% infection acquired immunity represented by the white line in (A.).

#### S1.4 Effect of Masking in Vaccinated and Unvaccinated Populations

We assess the effect of masking either only the vaccinated or unvaccinated populations for Type I and Type II infection spread types. Below are the trajectories of incidence per day for no masking and masking in only one subpopulation (see Figures S3 and S4).

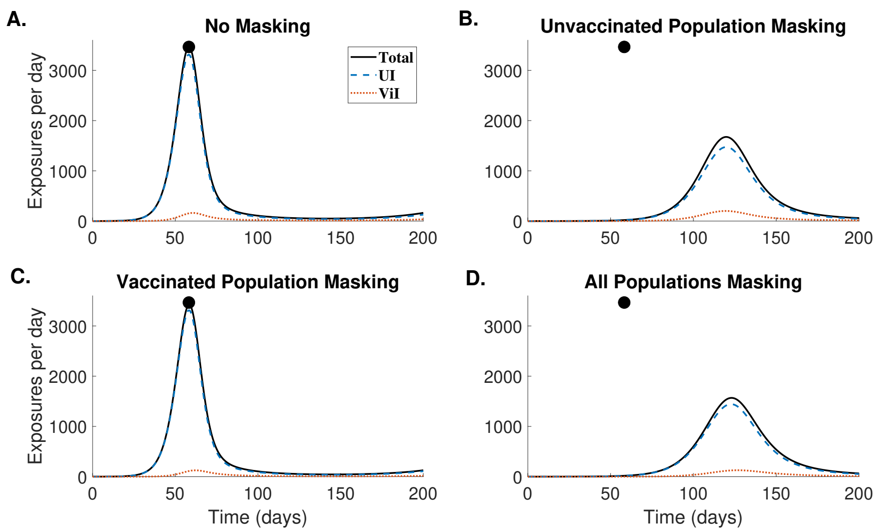


**Figure S3**: **During a Type I infection spread total incidence per day is reduced in cases where the unvaccinated population masks, but remains unchanged in the case that only the vaccinated population masks.** Total incidence per day per 100,000 (black) over 350 days generated from Model **G** with $\epsilon = 0.7$, $\phi = 0.1$, $\xi = 0.3$ (Type I infection spread), and parameters from Table S1. We assume masking reduces transmissions by 50% when both individuals involved in an exposure are masked or by 25% when one individual involved in an exposure is masked. Exposures are classified as involving only unvaccinated individuals, **UI** (blue), or involving at least 1 vaccinated individual, $\mathcal{I}_{\mathbf{U}\to\mathbf{V}}+\mathcal{I}_{\mathbf{V}\to\mathbf{U}}+\mathbf{VI}$ (red). **A.** No masking implemented. **B.** Masking is implemented in unvaccinated population only. **C.** Masking is implemented in the vaccinated population only. **D.** Masking is implemented in both unvaccinated and vaccinated populations. The black circle indicates the peak total incidence per day in the case of no masking.

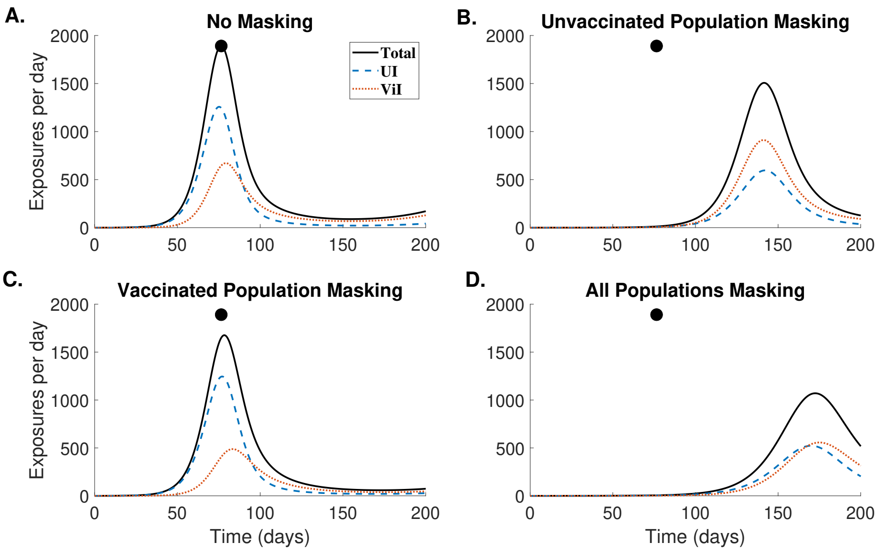


**Figure S4: During Type II infection spread total incidence per day is reduced most significantly in cases where both the unvaccinated and vaccinated populations mask, and reduced less significantly in the case that only the unvaccinated population or only the vaccinated population masks.** Total incidence per day per 100,000 (black) over 350 days generated from Model **G** with $\epsilon = 0.7$, $\phi = 0.5$, $\xi = 0.3$ (Type II infection spread), and parameters from Table S1. We assume masking reduces transmissions by 50% when both individuals involved in an exposure are masked or by 25% when one individual involved in an exposure is masked. Exposures are classified as involving only unvaccinated individuals, **UI** (blue), or involving at least 1 vaccinated individual, $\mathcal{I}_{\mathbf{U}\to\mathbf{V}}+\mathcal{I}_{\mathbf{V}\to\mathbf{U}}+$**VI** (red). **A.** No masking implemented. **B.** Masking is implemented in unvaccinated population only. **C.** Masking is implemented in the vaccinated population only. **D.** Masking is implemented in both unvaccinated and vaccinated populations. The black circle indicates the peak total incidence per day in the case of no masking.

Additionally, we test the effect of masking efficacy for each infection spread type. We very the reduction in transmission from 5% to 70% (low to high mask efficacy). The difference in maximum incidence per day and the difference in days to reach maximum incidence per day between the case of masking only in the unvaccinated sub-population versus masking in all sub-populations is given in Table S2.

#### S1.5 Effect of Increased Vaccine Transmission Blocking

Figure S5 demonstrates, using Model **G**, that as the effectiveness of a vaccine to block forward transmission increases, the parameter space where Type II and III infection spread occurs decreases. In other words, for vaccines that are highly effective

| | Difference in Peak Incidence per day (per 100,000) for Varying Masking Efficiencies | | | | | | | |
|---|---|---|---|---|---|---|---|---|
| Infection Spread Type | 5% | 10% | 20% | 30% | 40% | 50% | 60% | 70% |
| Type I | 5<br>(-0 days) | 10<br>(-0 days) | 22<br>(-0 days) | 40<br>(-0 dyas) | 66<br>(-1 days) | 107<br>(-3 days) | 174<br>(-11 days) | 280<br>(-57 days) |
| Type II | 48<br>(-0 days) | 111<br>(-1 days) | 194<br>(-2 days) | 302<br>(-5 days) | 437<br>(-13 days) | 596<br>(-31 days) | 794<br>(-76 days) | 1,070<br>(-203 days) |
| Type III | 40<br>(-2 days) | 82<br>(-4 days) | 173<br>(-12 days) | 270<br>(-25 days) | 372<br>(-48 days) | 484<br>(-88 days) | 633<br>(-163 days) | 998<br>(-318.5 days) |

**Table S2**: **The impact of the choice of masking strategy on maximum incidence per day for different infection spread types depends on the efficacy of the masking implementation.** Difference in the maximum incidence per day and days to reach maximum incidence per day between the case that only the unvaccinated sub-population versus all sub-populations implement masking. Maximum incidence per day generated from Model **G** with $\epsilon = 0.7$, $\phi = 0.1$, $\xi = 0.3$ (Type I infection spread), $\epsilon = 0.7$, $\phi = 0.5$, $\xi = 0.3$ (Type II infection spread), and $\epsilon = 0.7$, $\phi = 0.85$, $\xi = 0.3$ (Type III infection spread), and parameters from Table S1. We very the reduction in transmission from 5% to 70% (low to high mask efficacy).

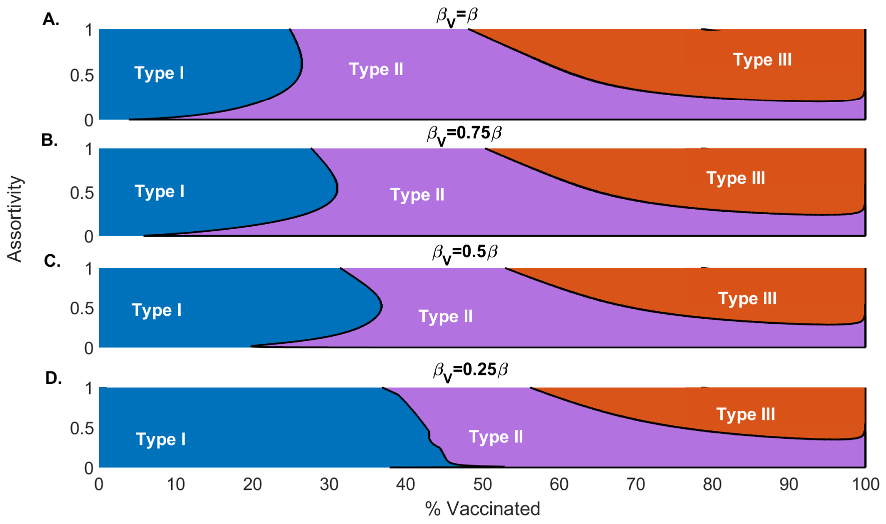


**Figure S5**: **In cases where vaccines are highly effective at reducing transmissions from vaccinated individuals, Type III infection spread is only possible at high vaccination rates and high population mixing.** Boundaries defining Types I, II, and III infection spread defined in the assortivity ($\epsilon$) and proportion vaccinated ($\phi$) parameter space for $\xi = 0.3$ and parameters given in Table S1. Vaccinated individuals have either (**A.**) the same infectivity, (**B.**) a 25% reduction in infectivity, (**C.**) a 50% reduction in infectivity , or (**D.**) a 75% reduction in infectivity compared to unvaccinated individuals.

at blocking transmission, we would only expect Type III infection spread at both high vaccination rates and high population mixing.

### S2: Colorado COVID-19 Tranmission Example

#### S2.1 Model C Equations

A system of Model **C** equations for one age-class $i$, where $i = \{0 \text{ to } 19, 20 \text{ to } 39, 40 \text{ to } 64, 64+\}$ is presented below.

$$
\begin{aligned}
\frac{dS_i^u}{dt} &= -\beta_I S_i^u(\mathcal{A}) - \beta S_i^u(\mathcal{B}) - \phi_i S_i^u + \frac{1}{\gamma}\lambda_A A_i^u \\
&\quad + \lambda_I\left(\frac{1}{\gamma}(1-d_{nhi}^u - h_i^u)I_i^u + \frac{1}{s_i}(1-d_{hi}^u)H_i^u\right) + \frac{1}{\tau_u}R_i^u \\
\frac{dE_i^u}{dt} &= \beta_I S_i^u(\mathcal{A}) + \beta S_i^u(\mathcal{B}) - \frac{1}{\alpha}E_i^u \\
\frac{dA_i^u}{dt} &= \frac{1-\rho_i}{\alpha}E_i^u - \frac{1}{\gamma}A_i^u \\
\frac{dI_i^u}{dt} &= \frac{\rho_i}{\alpha}E_i^u - \frac{1}{\gamma}I_i^u \\
\frac{dR_i^u}{dt} &= \frac{1}{\gamma}(1-\lambda_A)A_i^u + (1-\lambda_I)\left(\frac{1}{\gamma}(1-d_{nhi}^u - h_i^u)I_i^u + \frac{1}{s_i}(1-d_{hi}^u)H_i^u\right) \\
&\quad - \phi_i R_i^u - \frac{1}{\tau_u}R_i^u \\
\frac{dH_i^u}{dt} &= \frac{1}{\gamma}h_i^u I_i^u - \frac{1}{s_i}H_i^u \\
\frac{dV_i}{dt} &= \phi_i S_i^u + \phi_i R_i^u - \psi_i V_i - \frac{1}{\tau_v}V_i \\
\frac{dS_i^v}{dt} &= -\beta_I S_i^v(\mathcal{C}) - \beta S_i^v(\mathcal{D}) - \psi_i S_i^v + \frac{1}{\tau_b}R_i^v + \frac{1}{\tau_v}(V_i + B_i) \\
\frac{dE_i^v}{dt} &= \beta_I S_i^v(\mathcal{C}) + \beta S_i^v(\mathcal{D}) - \frac{1}{\alpha}E_i^v \\
\frac{dA_i^v}{dt} &= \frac{1-\rho_i}{\alpha}E_i^v - \frac{1}{\gamma}A_i^v \\
\frac{dI_i^v}{dt} &= \frac{\rho_i}{\alpha}E_i^v - \frac{1}{\gamma}I_i^v \\
\frac{dR_i^v}{dt} &= \frac{1}{\gamma}A_i^v + \frac{1}{\gamma}(1-d_{nhi}^v - h_i^v)I_i^v + \frac{1}{s_i}(1-d_{hi}^v)H_i^v - \psi_i R_i^u - \frac{1}{\tau_u}R_i^v \\
\frac{dH_i^v}{dt} &= \frac{1}{\gamma}h_i^v I_i^v - \frac{1}{s_i}H_i^v \\
\frac{dB_i^v}{dt} &= \psi_i S_i^v + \psi_i R_i^v + \psi_i V_i^v - \tau_v B_i^v \\
\frac{dD_i}{dt} &= \frac{1}{\gamma}d_{nhi}^u I_i^u + \frac{1}{\gamma}d_{nhi}^v I_i^v + \frac{1}{s_i}d_{hi}^u H_i^u + \frac{1}{s_i}d_{hi}^v H_i^v,
\end{aligned}
\tag{S2}
$$

where

$$\mathcal{A} = \sum_{j=1}^{4} \left[ (1-\epsilon)\frac{I_j^u}{N^u} + \epsilon \left( \frac{I_j^u + I_j^v}{N} \right) \right], \qquad \mathcal{B} = \sum_{j=1}^{4} \left[ (1-\epsilon)\frac{A_j^u}{N^u} + \epsilon \left( \frac{A_j^u + A_j^v}{N} \right) \right],$$

$$\mathcal{C} = \sum_{j=1}^{4} \left[ (1-\epsilon)\frac{I_j^v}{N^v} + \epsilon \left( \frac{I_j^u + I_j^v}{N} \right) \right], \qquad \mathcal{D} = \sum_{j=1}^{4} \left[ (1-\epsilon)\frac{A_j^v}{N^v} + \epsilon \left( \frac{A_j^u + A_j^v}{N} \right) \right].$$

Note that capital letters refer to disease status compartments and $u$ or $v$ superscripts indicate vaccination status (unvaccinated or vaccinated, respectively).

The formula for **UI**, $\mathcal{I}_{\mathbf{U}\to\mathbf{V}}$, $\mathcal{I}_{\mathbf{V}\to\mathbf{U}}$, and **VI** for a single age-class $i$, where $i = \{0 \text{ to } 19, 20 \text{ to } 39,\ 40 \text{ to } 64, 64+\}$ are presented below.

$$\mathcal{I}_{\mathbf{U}\to\mathbf{U}}(t) := \beta_I S_i^u \sum_{j=1}^{4} \left[ (1-\epsilon)\frac{I_j^u}{N^u} + \epsilon \left( \frac{I_j^u}{N} \right) \right] + \beta S_i^u \sum_{j=1}^{4} \left[ (1-\epsilon)\frac{A_j^u}{N^u} + \epsilon \left( \frac{A_j^u}{N} \right) \right] \tag{S3}$$

$$\mathcal{I}_{\mathbf{U}\to\mathbf{V}}(t) := \beta_I S_i^v \sum_{j=1}^{4} \epsilon \left( \frac{I_j^u}{N} \right) + \beta S_i^v \sum_{j=1}^{4} \epsilon \left( \frac{A_j^u}{N} \right) \tag{S4}$$

$$\mathcal{I}_{\mathbf{V}\to\mathbf{U}}(t) := \beta_I S_i^u \sum_{j=1}^{4} \left[ \epsilon \left( \frac{I_j^v}{N} \right) \right] + \beta S_i^u \sum_{j=1}^{4} \left[ \epsilon \left( \frac{A_j^v}{N} \right) \right] \tag{S5}$$

$$\mathcal{I}_{\mathbf{V}\to\mathbf{V}}(t) := \beta_I S_i^v \sum_{j=1}^{4} \left[ (1-\epsilon)\frac{I_j^v}{N^v} + \epsilon \left( \frac{I_j^v}{N} \right) \right] + \beta S_i^v \sum_{j=1}^{4} \left[ (1-\epsilon)\frac{A_j^v}{N^v} + \epsilon \left( \frac{A_j^v}{N} \right) \right]. \tag{S6}$$

For combined age-classes, $\mathcal{I}_{\mathbf{U}\to\mathbf{U}} = \sum_j \mathcal{I}_{\mathbf{U}\to\mathbf{U}j}$, $\mathcal{I}_{\mathbf{U}\to\mathbf{V}} = \sum_j \mathcal{I}_{\mathbf{U}\to\mathbf{V}j}$, $\mathcal{I}_{\mathbf{V}\to\mathbf{U}} = \sum_j \mathcal{I}_{\mathbf{V}\to\mathbf{U}j}$, and $\mathcal{I}_{\mathbf{V}\to\mathbf{V}} = \sum_j \mathcal{I}_{\mathbf{V}\to\mathbf{V}j}$.

#### S2.3 Model C Parameters

The table below presents the fixed values of parameters used in data-fitting and simulations of Model **C**.

| parameter | description | unit | value |
|---|---|---|---|
| $\beta$ | asymptomatic transmission rate | per day | fit |
| $\beta_I$ | infectious transmission rate | days | $1.395 \times \beta$ |
| $\alpha$ | incubation period | days | 2.5 |
| $\gamma$ | length of infection | days | 5 |
| $\tau_U$ | length of protection from infection acquired immunity | days | 365 |
| $\tau_b$ | length of protection from vaccine acquired immunity | days | 213 |
| $\tau_V$ | length of protection from infection acquired immunity after vaccination | days | 213 |
| $N_j$ | population of age class $j$ | people | $[1,411,161\ \ 1,697,671\ \ 1,818,147\ \ 878,522]$ |
| $\epsilon$ | assortivity between unvaccinated and vaccinated populations | | $[0,1]$ |
| $\lambda_A$ | fraction of unvaccinated asymptomatics without acquired immunity after infection | | 0.15 |
| $\lambda_I$ | fraction of unvaccinated infectious without acquired immunity after infection | | 0.075 |
| $d_{nhj}$ | proportion of unvaccinated non-hospitalized deaths for age class $j$ | | $[1.3 \times 10^{-5}\ \ 8.2 \times 10^{-5}\ \ 6.2 \times 10^{-4}\ \ 2.8 \times 10^{-2}]$ |
| $d_{hj}$ | proportion of hospitalized deaths for age class $j$ | | $[5.5 \times 10^{-3}\ \ 1.5 \times 10^{-2}\ \ 5.8 \times 10^{-2}\ \ 0.16]$ |
| $h_j$ | proportion of unvaccinated hospitalizations for age class $j$ | | $[0.039\ \ 0.062\ \ 0.086\ \ 0.1343]$ |
| $rho_j$ | proportion of infectious cases for age class $j$ | | $[0.11\ \ 0.36\ \ 0.56\ \ 0.78]$ |
| $phi_j$ | vaccination rate for age class $j$ | per day | from data |
| $Phi_j$ | booster rate for age class $j$ | per day | from data |
| $s_j$ | length of hospitalization for age class $j$ | days | from data |

**Table S3**: Parameter values used in data-fitting and simulations of Model **C**. *Hospitalization rate was changed to account for new variants by applying the following multipliers for every age class: 1.2243 for B.1.1.7 variant on 2/15/2021, 1.5598 on 5/15/21, 1.681 for Delta variant on 7/15/2021, and 0.5 for Omicron on 01/01/2022.
